# County by county estimation of possible deaths averted, infections averted, and cost savings of overdose prevention centers in the United States from July 2019-June 2025

**DOI:** 10.1101/2024.09.21.24314113

**Authors:** Katie Bailey, Rohith Palli, Mike Sportiello

## Abstract

Overdose prevention centers (OPCs), also known as safer injection facilities (SIFs) or supervised consumption sites (SCSs), are a service that allows people to obtain sterile supplies to use their own drugs in a safe environment that is monitored by professionals trained in overdose response and harm reduction. Though still illegal under federal law in the U.S. (“Maintaining Drug-Involved Premises”; “United States v. Safehouse”, 2021), many legal or quasi-legal (including some in the U.S.) OPCs have operated since the 1980s. Using CDC overdose death estimates and U.S. Census Bureau population data, we developed a retrospective, scenario-based modeling tool, estimating that 1,329-5,316 deaths could have been averted between July 2019 and June 2025 if 1 out of 100-400 injections had taken place at an OPC. Under these assumptions, 26-105 HIV transmissions and 5,723-22,892 Hepatitis C transmissions could have been averted. Factoring in both discounted lifetime costs to treat those infections as well as the statistical value of deaths averted, $11.9-47.7 billion could have been saved. These data support OPCs as one viable public health intervention to avert deaths and avoid infections. We created a web application that allows users to explore county-level model results and alter model assumptions, including variables like the percent of injections that occur at an OPC.

## Introduction

Overdose prevention centers (OPCs), also known as supervised consumption sites (SCSs) or safer injection facilities (SIFs) when designed primarily for people who inject drugs (PWID), offer a monitored setting where people can use pre-obtained drugs with access to sterile supplies, staff trained in overdose response, and referrals to medical, behavioral health, and social services. Many legal or quasi-legal OPCs have operated in multiple countries since the 1980s, including some in the U.S., although they are not clearly authorized under U.S. federal law (Beletsky et al., 2018; “Maintaining Drug-Involved Premises”; “United States v. Safehouse”, 2021). Across these settings, overdoses have been managed on-site, with no deaths reported in supervised consumption spaces, in large part due to rapid intervention with naloxone, an opioid overdose reversal agent, and respiratory support such as supplemental oxygen or bag-mask ventilation (Suen et al., 2023).

Research from Canada suggests that OPCs can reduce overdose-related harms, including mortality (Gariepy et al., Sites de consommation supervisee et mortalite par surdose a l’echelle de la population : revue systematique des donnees recentes, 2016-2024./2025). For example, a study of Vancouver’s first OPC found a 35% reduction in overdose deaths in the surrounding area compared to smaller declines in the rest of the city (Marshall et al., 2011). In Toronto, spatial analyses estimated that approximately two overdose deaths per 100,000 people were averted annually in areas closest to OPCs (Rammohan et al., 2024). Additional analyses from Ontario found more favorable overdose mortality trends and fewer overdose-related adverse outcomes in areas with greater OPC presence or capacity (Panagiotoglou & Lim, 2022; T. Robinson et al., 2024; Yeung et al., 2023), and individual- and population-level studies from Vancouver and Alberta have similarly suggested that greater OPC engagement is associated with lower overdose mortality and related harms (Kennedy et al., 2019; Marshall et al., 2021).

Findings, however, have not been uniformly favorable. A controlled interrupted time-series study from Ontario and a linked cohort analysis from Toronto reported null, mixed, or limited short-term associations between OPC availability and opioid-related mortality, acute care use, or other health outcomes at the population level (Day et al., 2026; Tessa Robinson et al., 2024). These mixed findings likely reflect differences in study design, follow-up time, geographic scale, service intensity, and the difficulty of isolating OPC effects amid a rapidly evolving overdose crisis (Gariepy et al., Sites de consommation supervisee et mortalite par surdose a l’echelle de la population : revue systematique des donnees recentes, 2016-2024./2025). In the U.S., evidence remains more limited due to legal restrictions, but includes observational studies of an unsanctioned OPC (Kral et al., 2020), the first two publicly recognized OPCs in New York City (Harocopos et al., 2022), and a sanctioned San Francisco-based OPC that has since closed (Suen et al., 2023), with each reporting high utilization, numerous overdose events managed on-site, and no observed fatal overdoses within supervised settings.

Modeling studies have also projected meaningful public health and economic benefits of OPC implementation in specific Canadian and U.S. cities, including estimated reductions in overdose deaths, infectious disease transmission, ambulance and emergency department utilization, and health care costs, with resulting favorable cost-effectiveness estimates (Andresen & Boyd, 2010; Bayoumi & Zaric, 2008; Irwin et al., 2016; Irwin et al., 2017; Jozaghi et al., 2013). A recent systematic review of economic evaluations similarly concluded that OPC implementation was consistently associated with favorable economic outcomes across North American settings (Behrends et al., 2024). Importantly, however, OPCs may be better aligned with some patterns of drug use and overdose risk than others. Most evidence on OPC outcomes has focused on opioid-related overdose and supervised injection drug use, while overdose risk in the U.S. increasingly reflects non-injection routes of use, including smoking (Ciccarone et al., 2024; Eger et al., 2024; Karandinos et al., 2024; Kral et al., 2021), as well as growing stimulant involvement (Tanz et al., 2025). Although some OPCs are adapting to include ventilated spaces for inhalation (OnPoint, 2025), these services have not traditionally been part of the OPC model. As a result, the impact of OPCs likely varies depending on local patterns of drug use and the extent to which site services align with the needs of people at highest risk of overdose.

We build on previous literature to explore the potential impact of OPC implementation across U.S. communities while accounting for variation in local conditions and implementation assumptions (Harocopos et al., 2022; Kral et al., 2020). To do so, we developed a retrospective, scenario-based modeling tool to estimate a range of possible public health and economic outcomes if OPCs had been implemented across U.S. counties between July 2019 and June 2025. Specifically, we estimate potential deaths, HIV and Hepatitis C virus (HCV) infections, and downstream economic burden averted under varying assumptions about key implementation and utilization parameters, such as frequency of OPC use among people who inject drugs (PWID). Because the model is primarily based on assumptions related to supervised injection of opioids, these estimates are likely most applicable to settings where opioid injection was a major driver of overdose risk. Rather than estimating a single nationwide causal effect of OPCs, we developed an interactive web application that enables policymakers, advocates, and other stakeholders to explore how OPC implementation might have influenced county-level public health outcomes. We further illustrate the application of this tool through three county-level case studies, highlighting how estimated outcomes vary across diverse settings and under different implementation assumptions.

## Methods

Provisional 12-month ending drug overdose deaths by county were obtained from the National Center for Health Statistics for June of each year from 2020 through 2025 (Ahmad et al., 2026); each estimate represents deaths occurring over the preceding 12 months (July–June), yielding non-overlapping annual periods. Population and age data were downloaded from the U.S. Census Bureau. For this analysis, we assumed 1.5% of adults were PWID per previous estimates (Bradley et al., 2026; Bradley et al., 2023). For each PWID, we estimated 508.8 injections per year, or just under 1.5 injections based on prior estimates (Bradley et al., 2026; Bradley et al., 2023).

### Deaths averted

Deaths averted were calculated by assuming the total number of injections stayed constant. Secondly, we assumed that deaths cannot occur after an injection in the OPC, as no reported overdose death has ever occurred in an OPC. The *injections that would occur at an OPC* were calculated by modeling 0.25%, 0.5%, 0.75%, or 1% of injections taking place at the OPC. This led to the following calculation:

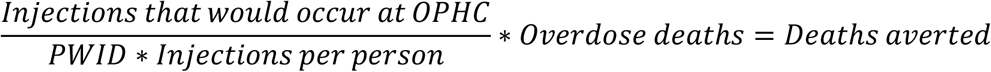

With the fraction on the left simplifying to the frequency of total injections in the OPC, multiplied by the number of deaths. If all the injections took place at the OPC, the leftmost fraction would simplify to 1 and all deaths would be averted. If none did, the whole left term would simplify to 0 and no deaths would be averted.

### HCV transmissions averted

HCV is a highly transmissible virus in the setting of shared needle use (Pouget et al., 2012), leading to significant morbidity, mortality, and health expenditures (Stepanova & Younossi, 2017). While curative therapy now exists, it is expensive and requires weeks-long adherence, which can be difficult for many PWID as they are disproportionately housing-insecure and have persistent barriers to healthcare engagement (Austin et al., 2022; Kimball et al., 2024). Similar to deaths averted, due to the supply of sterile drug use equipment including needles and cottons, transmission of HCV has never been documented at an OPC. Natural incidence of HCV within PWID varies widely, with study estimates varying from 0.51 to 47 per 100 person years (Gutfraind et al., 2015). For this analysis, we used a lower-end estimate of 10 per 100 person years (Gutfraind et al., 2015), leading to the following equation:

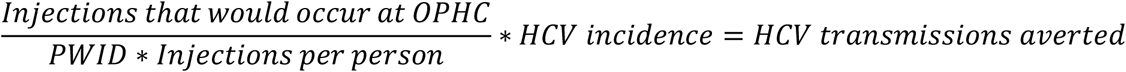

### HIV transmissions averted

Though the vast majority of HIV is not transmitted through injection drug use, it is still a meaningful target of HIV transmission prevention efforts in the U.S. (Centers for Disease & Prevention, 2024). Others have calculated the incidence of HIV transmission in groups of PWID, stratifying their use as low, medium, and high risk depending on factors including how often they share needles with people they know and alternatively with strangers (Jarlais et al., 2022). Using these incidences, we calculated HIV transmissions averted with the following equations:

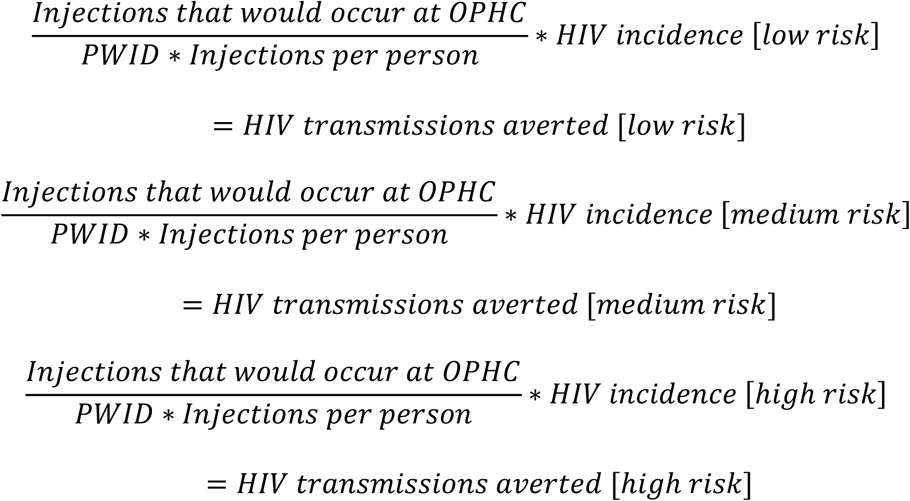

### Costs averted

Substantial health care dollars are spent treating preventable diseases. Infectious diseases can be prevented with appropriate education and low-cost materials. Others have estimated the discounted lifetime cost of an HIV infection to be $448,095 after adjusting for inflation (Schackman et al., 2015). We calculated the discounted lifetime cost of an HIV infection with the following equation:

$448,095 ∗ *averted HIV transmission* = *Discounted cost of an averted HIV infection*

Similar work has been done to calculate the discounted lifetime cost of HCV, calculated to $339,968 (after adjusting for inflation) (Liu et al., 2016).

$339,968 ∗ *averted HIV transmission* = *Discounted cost of an averted HCV infection*

Finally, we calculated the total economic costs of the lives lost to overdose, using

$7.5 million as the statistical value of life, consistent with conservative estimates used by

U.S. federal agencies (Federal Emergency Management), giving the following equation:

$7,500,000 ∗ *Averted deaths = Statistical costs of deaths averted*

In this work, total costs would include adding up both the discounted costs of HIV and HCV infections as well as statistical costs of deaths averted:

*Discounted infectious costs + Statistical of daths averted = Total costs averted*

Of note, total costs averted do not include indirect benefits, such as potential reductions in HIV and HCV transmission outside of the OPC or decreased emergency service utilization associated with on-site overdose management.

All analysis was performed using R and can be found publicly available here: https://github.com/sportiellomike/OPC-analysis-code.

### Web application

A free web application (“OPC Data Explorer”) was created using the Shiny infrastructure and written in R (Alboukadel, 2023; Aron et al., 2025; Edzer, 2018; Edzer & Roger, 2023; Elio, 2025; Garnier et al., 2024; Gregory et al., 2023; Hadley, 2007; Hadley, Lionel, et al., 2025; Hadley et al., 2019; Hadley, Thomas Lin, et al., 2025; Jeffrey, 2024; Jeroen, 2025; Kyle & Matt, 2025; Mark et al., 2017; Paolo Di, 2024; Scott et al., 2023; Sergei, 2024; Team, 2025; Teun van den, 2025; Tyson et al., 2025; Winston, 2021; Winston et al., 2024; Yihui et al., 2024). All code for the data explorer web application is free and open source and can be found here: https://github.com/sportiellomike/OPC-data-explorer. The most up to date version of the web application itself can be found here: https://sportiellomike.shinyapps.io/OPC-data-explorer/. Minimal assistance in coding was provided by generative artificial intelligence (AI) using claude-3.5-sonnet. All additions provided by AI were manually checked at the time of coding for both function and syntax. Generative AI was not utilized to create or manipulate any figure.

## Results

### Analytic results

Estimates of potential deaths averted are shaped by assumptions regarding the percentage of injections that occur at an OPC (Table 1). At the low end, assuming 0.25% of injections occurred at OPCs across U.S. counties, (1 injection out of 400, or one injection every 287 days [400/(508.8/365)]) per PWID, an estimated 1,329 deaths may have been averted nationally. At the higher end of 1% of injections occurring at an OPC, 5,316 deaths may have been averted.

**Table 1:** Summary table of estimated overdose deaths, infectious diseases, and downstream economic burden averted under modeled Overdose Prevention Centers implementation across U.S. counties, July 2019-June 2025

| Percent of injections occurring at OPC | 0.25% | 0.5% | 0.75% | 1% |
| --- | --- | --- | --- | --- |
| **Deaths averted** | 1328.91 | 2657.82 | 3986.73 | 5315.64 |
| **HCV infections averted** | 5722.9 | 11445.98 | 17168.98 | 22891.97 |
| **HIV-low infections averted** | 11.45 | 22.89 | 34.34 | 45.78 |
| **HIV-medium infections averted** | 26.33 | 52.65 | 78.98 | 105.30 |
| **HIV-high infections averted** | 32.05 | 64.10 | 96.15 | 128.20 |
| **Discounted lifetime cost infections (billions of dollars)** | 1.96 | 3.91 | 5.87 | 7.83 |
| **Cost of life (billions of dollars)** | 9.97 | 19.93 | 29.90 | 39.87 |
| **Total costs (billions of dollars)** | 11.92 | 23.85 | 35.77 | 47.70 |

Estimates of HCV infections averted range from 5,723 to just under 22,892 (Table 1), and dozens of HIV transmissions may have been averted (Table 1). Across all OPC utilization scenarios, estimated downstream economic burden potentially averted was in the range of billions to tens of billions of dollars.

Even under more limited implementation scenarios, such as restricting OPCs to the top 50 counties with the largest estimated populations of PWID (Table 2) or those with the 50 highest rates of overdose deaths per capita (Table 3), projected impacts remained substantial. A list of these counties can be found in supplementary Tables 1 and 2 respectively.

**Table 2:** Estimated overdose deaths, infectious diseases, and downstream economic burden averted under modeled Overdose Prevention Centers implementation across the 50 U.S. counties with the greatest number of people who inject drugs per capita, July 2019-June 2025

| Percent of injections occurring at OPC | 0.25% | 0.5% | 0.75% | 1% |
| --- | --- | --- | --- | --- |
| **Deaths averted** | 402.49 | 804.97 | 1207.46 | 1609.94 |
| **HCV infections averted** | 1738.93 | 3477.85 | 5216.78 | 6955.71 |
| **HIV-low infections averted** | 3.48 | 6.96 | 10.43 | 13.91 |
| **HIV-medium infections averted** | 8.00 | 16.00 | 24.00 | 32.00 |
| **HIV-high infections averted** | 9.74 | 19.48 | 29.21 | 38.95 |
| **Discounted lifetime cost of infections (billions of dollars)** | 0.59 | 1.19 | 1.78 | 2.38 |
| **Cost of life (billions of dollars)** | 3.02 | 6.04 | 9.06 | 12.07 |
| **Total costs (billions of dollars)** | 0.59 | 1.19 | 1.78 | 2.38 |

**Table 3:** Estimated overdose deaths, infectious diseases, and downstream economic burden averted under modeled Overdose Prevention Center implementation across 50 U.S. counties with the highest rates of overdose death per capita, July 2019-June 2025

| Percent of injections occurring at OPC | 0.25% | 0.5% | 0.75 | 1% |
| --- | --- | --- | --- | --- |
| **Deaths averted** | 52.77 | 105.54 | 158.31 | 211.08 |
| **HCV infections averted** | 83.82 | 167.64 | 251.46 | 335.28 |
| **HIV-low infections averted** | 0.17 | 0.34 | 0.50 | 0.67 |
| **HIV-medium infections averted** | 0.39 | 0.77 | 1.16 | 1.54 |
| **HIV-high infections averted** | 0.47 | 0.94 | 1.41 | 1.88 |
| **Discounted lifetime cost of infections (millions of dollars)** | 0.03 | 0.06 | 0.09 | 0.11 |
| Cost of life (billions of dollars) | 0.40 | 0.79 | 1.19 | 1.58 |
| Total costs (billions of dollars) | 0.03 | 0.06 | 0.09 | 0.11 |

### Web application

The open-source OPC Data Explorer interactive web application was constructed for users to investigate estimates in individual counties and create color-coded maps and tables with additional parameters not reported here (discounted costs broken down between HIV and HCV, for example) (Figure 1). The most up-to-date version can always be found at: https://sportiellomike.shinyapps.io/OPC-data-explorer/. Users can dive into the data and share downloadable figures and analytic tables.

**Figure 1:**
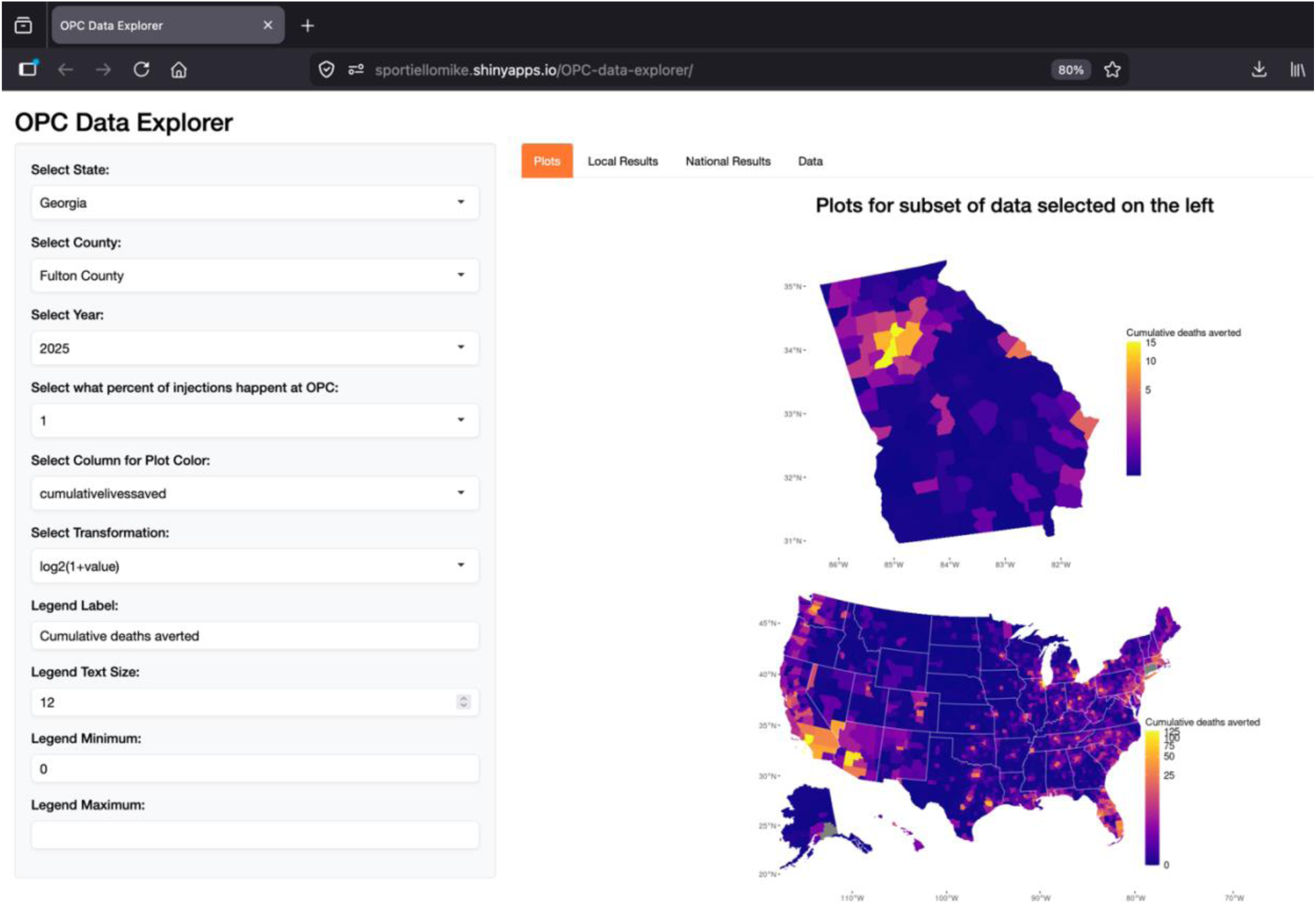
Overdose Prevention Center Data Explorer Web Application

### Case studies

To illustrate variations in estimates of how OPC implementation may have impacted overdose, infectious disease, and economic burden across local contexts, we present estimates for three counties selected to reflect different epidemiologic and geographic settings. These examples are intended to demonstrate how the modeling tool may support local interpretation and planning.

### Fulton County, Georgia

Georgia’s Fulton County is the most populous in the state and includes most of the city of Atlanta and its surrounding suburbs. With approximately one million residents, Fulton County has one of the highest burdens of both overdose mortality and HIV in the country (Centers for Disease & Prevention, 2024), making it a useful example of a large urban county where OPC implementation could have substantial impact. The county is one of the priority jurisdictions in the Ending the HIV Epidemic initiative, with an estimated 2022 HIV incidence of 23.1 per 100,000 people (U. S. Department of Health & Human Services, 2026a). Similar to national trends and the case studies described below, deaths from inhalation and stimulant use continue to rise in Georgia (Georgia Department of Public Health, 2025a, 2025b).

Between July 2019 and June 2025, 1,519 Fulton County residents died from overdose (Ahmad et al., 2026). Under modeled OPC implementation scenarios, we estimated that 4–16 overdose deaths and 19–75 HCV infections may have been averted, with minimal impact on HIV transmission (Table 4). The estimated economic value associated with harms averted ranged from $35–140 million.

**Table 4:** County-specific estimates of overdose deaths, infectious diseases, and downstream economic burden averted under modeled Overdose Prevention Center implementation for select counties, July 2019–June 2025^a^

|  | Fulton County,<br>GA | Monroe County,<br>NY | San Diego County,<br>CA |
| --- | --- | --- | --- |
| **Real world overdose deaths** | 1519 | 1705 | 4940 |
| **Deaths averted** | 4-16 | 4-17 | 12-49 |
| **HCV averted** | 19-75 | 13-54 | 58-233 |
| **HIV-low averted** | 0 | 0 | 0 |
| **HIV-medium averted** | 0 | 0 | 0-1 |
| **HIV-high averted** | 0 | 0 | 0-1 |
| **Discounted lifetime cost infections (millions of dollars)** | 6-26 | 5-20 | 20-80 |
| **Cost of life (millions of dollars)** | 28-114 | 32-128 | 93-371 |
| **Total costs (millions of dollars)** | 35-140 | 37-146 | 113-450 |

<sup>a</sup>Data listed as ranges within each cell ranging from estimate at 1/400 injections to 1/100 injections.

Despite being located in a non-Medicaid expansion state with comparatively lower public health investment, Fulton County has developed a range of services to address overdose risk (Georgia Overdose Prevention, 2026). However, the substantial burden of overdose mortality indicates that existing services may not fully meet population need. In this context, the modeled estimates suggest that even low levels of OPC utilization could yield measurable reductions in overdose deaths and HCV infections. At the same time, the modest population-level impact highlights that higher levels of OPC coverage would likely be necessary to meaningfully reduce overall overdose mortality. Because the model assumes a uniform prevalence of injection drug use (1.5% of the adult population), these estimates may be conservative for Southern counties like Fulton, where the population of people who inject drugs may be larger (Altekruse et al., 2020; Bradley et al., 2026).

### Monroe County, New York

New York’s Monroe County is the ninth most populous in the state with approximately 750,000 residents. The county covers a mix of urban, suburban, and rural areas with residents of the city of Rochester, the state’s fourth-largest city, making up approximately 27% of the total population. Although Monroe County includes a major urban center, many overdose deaths occur outside the urban core (New York State Department of Health, 2026). While overdose death rates have declined more than national trends in recent years, the local drug supply has shifted, with increasing involvement of stimulants and xylazine (Granger & Hartman, 2025).

During the study period, 1,705 Monroe County residents died from overdose (Table 4). Under modeled OPC implementation scenarios, we estimated that between 4-17 overdose deaths may have been averted in Monroe County between July 2019 and June 2025, and 13-54 HCV infections may have been averted (Table 4). Minimal HIV infections are projected to have been averted by any scenario. Projected downstream economic burden potentially averted ranged from $37-$146 million.

New York State has been a national leader in expanding harm reduction services, including broad access to sterile syringes and the implementation of sanctioned OPCs in New York City (New York State Department of Health, 2024). In Monroe County, these efforts are reflected in a relatively robust network of public and non-profit syringe service programs and naloxone distribution across both fixed and mobile settings (Monroe County Department of Public Health, 2026; New York State Department of Health, 2024). However, the distribution of overdose deaths outside the urban core highlights potential gaps in reach, particularly in suburban and rural areas. In this context, OPC implementation in Monroe County would likely require careful consideration of geographic need and accessibility, with potential value in complementing existing services by engaging populations not consistently reached through current approaches.

### San Diego County, California

San Diego County, California, is a large urban county whose proximity to the U.S.–Mexico border shapes local drug markets and overdose risk. As part of the San Diego– Imperial Valley High Intensity Drug Trafficking Area (Office of National Drug Control, 2025), the county has long been situated within a major corridor for drug trafficking while also facing substantial burden related to overdose and infectious disease. It is also a federal priority jurisdiction in the Ending the HIV Epidemic initiative, with an estimated HIV incidence of 14.4 per 100,000 people in 2022 (U. S. Department of Health & Human Services, 2026b), and more than 55,000 residents with a history of HCV infection (Cheema et al., 2024).

During the span of this analysis, 4,940 San Diego County residents died from overdose. Under modeled OPC implementation scenarios, we estimated that between 12-49 overdose deaths may have been averted in San Diego County between July 2019 and June 2025. Over the same period, up to 1 HIV infection and 58-233 HCV infections may have been averted. Projected downstream economic burden potentially averted ranged from $113-$450 million.

Situated in a state with comparatively supportive harm reduction policies, San Diego County’s harm reduction infrastructure expanded during the study period and includes multiple syringe services programs offered at fixed sites and via mobile outreach and widespread naloxone distribution (Bailey et al., 2024; County of San Diego Health & Human Services Agency, 2024). In this context, the modeled estimates suggest that OPC implementation could provide additional reductions in overdose deaths beyond those achieved through existing services and support local HCV prevention goals (Cheema et al., 2024; U. S. Department of Health & Human Services, 2026b).

## Discussion

While it is difficult to estimate the potential use of OPCs among PWID, this study supplies a framework to assist in estimating their impact on deaths, infections, and downstream economic burden. Even at very low rates of use, assuming one out of every 400 injections occurs at an OPC, hundreds of deaths and thousands of chronic infections may have been averted over the period studied. This study also modeled OPC implementation scenarios in the 50 counties with the most PWID and the 50 counties with the highest per capita overdose deaths, a potentially more realistic and cost-effective strategy that we estimate would have had a substantial positive impact on deaths, infections, and economic burden. We highlighted three counties as brief case studies to illustrate how this model can be applied in diverse local contexts

Our study has several limitations. Importantly, this analysis does not include secondary benefits of OPCs, like the potential for participants to voluntarily engage in treatment programs through referrals from OPC staff (Wood et al., 2007). It also does not include potential benefits of safer drug use education, such as decreasing the rates of needle sharing (Kerr et al., 2005). Lastly, this analysis does not include the potential OPC benefits of connection to secondary social supports, including housing, food, healthcare, or other programs. This analysis does not include any secondary detriments of OPCs, though it should be noted that most of those of concern (e.g., increase in overdoses, increase in new-onset illicit injection drug use) have not been borne out by studies that that have actually measured these important issues of concern (Kerr et al., 2005; Wood et al., 2004).

Another important aspect not included in this analysis is the cost to implement and sustain OPCs, which would require capital and start-up costs, staff salaries, ongoing supplies, and the costs of maintaining these institutions over time. Another drawback of this analysis is that its estimations are retrospective rather than projected: we do not make estimates regarding the impact of future OPCs, just that OPCs could have averted past adverse outcomes.

Another limitation of this study is the uncertainty in estimating the proportion of injections that would occur within OPCs if implemented. OPC utilization is likely to vary across settings based on local infrastructure, service availability, and community context, as well as characteristics of OPC design and implementation. In addition, the estimated impact of OPCs on infectious disease transmission may depend on the existing harm reduction environment; in settings with high coverage of sterile syringe access and other services, the marginal reductions in HIV and HCV transmission attributable to OPCs may be smaller. Drug use patterns vary widely across substances, populations, settings, and interactions with healthcare and law enforcement, even within the same city or neighborhood, and those who use OPCs may also be motivated to use other harm reduction resources. A rate of 1 in 400 injections occurring at an OPC would likely arise from a skewed distribution of use, with some individuals using the OPC frequently and many using it rarely. As no previous study has estimated the likelihood of OPC uptake per injection, we presented here a conservative and broad range of 0.25%-1% of injections occurring at an OPC. Another source of uncertainty is the estimate of injection rates in the communities without OPCs, which continues to be under debate in the literature and likely varies across time, neighborhoods, drug supplies, and more. Better objective data is needed to make these estimates precise.

Relatedly, the feasibility of OPCs has been called into question, especially for more rural counties where people live less densely: would those who would use the OPC be able to or desire to travel so far to an OPC? Would there be enough use of the OPC to justify the cost to set up and run it? As we are not aware of any objective data to help guide the estimates to these questions, we opted to treat more rural counties identically to the denser counties, but as we acknowledge, county density likely plays a large role in implementation decisions. As such, we presented model estimates given OPC implementation in more realistic scenarios based on population size and overdose burden.

The density critique stems from a traditional but potentially limiting view of OPCs as standalone structures with dedicated supplies and staff that are independently run. However, other health care delivery models demonstrate that OPCs need not be fixed spaces and may be deployed along a spectrum of permanence and independence. In some communities, especially the densest, it may make sense for OPCs to exist as standalone, physical structures. However, in less dense communities, OPC-type services could exist within rural hospital settings, similar to how some rural specialist practices operate. In even less dense settings, a mobile OPC similar to mobile dentistry and mobile mammograms may be a solution (Gao et al., 2019; Reuben et al., 2002). Ultimately, our data reflect not a strict analysis of a pre- and post-intervention, but the large impact minor reductions in relatively unsafe drug use can have at scale.

As mentioned previously, inhalation overdose deaths continue to rise. This trend likely reflects multiple factors, including the increasing presence of fentanyl in the drug supply and shifts in routes of administration away from injection toward inhalation, facilitated by the distribution of safer smoking supplies (Eger et al., 2024). Similarly, deaths associated with stimulant use, also often via inhalation, continue to rise (Friedman & Shover, 2023). Future OPCs should be designed to support those who smoke drugs and those who use stimulants. Relative to opioid overdoses, stimulant-related overdoses involve different clinical presentations and do not have a direct pharmacologic reversal agent such as naloxone. As a result, OPC models that have historically focused on opioid overdose response may require adaptation to effectively address stimulant and polysubstance-related harms. We were not able to examine outcomes based on route of administration (injection, inhalation, other) as this information is not codified in the CDC overdose death dataset.

In the end, with these caveats in mind, we acknowledge that the total mortality and morbidity of the ongoing overdose epidemic is difficult to quantify objectively, and that there is often large degree of confounding in computing the effect of interventions. What we proposed here is that, based on our modeling approach, OPC implementation could have contributed to reductions in overdose mortality and morbidity, with effects that remain meaningful even under conservative assumptions. The exact number of these estimates could and should be better estimated at the local level and measured objectively once an intervention is in place.

We invite stakeholders to use our interactive web tool to inform policy discussions about whether the potential benefits of OPCs outweigh the costs in their communities, recognizing that these benefits are likely to vary across settings.

## Conclusion

Overall, this analysis, facilitated by our interactive, web-based modeling tool, suggests that OPCs could contribute to reductions in overdose mortality and HCV transmission, with impacts that vary across local contexts. By enabling stakeholders to estimate potential effects in their own communities, this work highlights both the potential value of OPCs and the importance of context-specific implementation. At the same time, these findings underscore the need for additional empirical research to refine key assumptions and improve the precision of future estimates.

## Data Availability

All data produced in the present study are available upon reasonable request to the authors.

https://www.cdc.gov/nchs/index.html

https://data.census.gov/

## Acknowledgements

MS was supported by NIH grant T32GM152318 during some of the time of this work. KB is supported by NIDA grant T32DA023356.

---

**Supplementary table 1:** Counties with highest number of PWID

| Counties with highest number of PWID |
| --- |
| California-Los Angeles County |
| Illinois-Cook County |
| Texas-Harris County |
| Arizona-Maricopa County |
| California-San Diego County |
| California-Orange County |
| Florida-Miami-Dade County |
| New York-Kings County |
| Texas-Dallas County |
| New York-Queens County |
| California-Riverside County |
| Washington-King County |
| Nevada-Clark County |
| California-San Bernardino County |
| Texas-Tarrant County |
| Florida-Broward County |
| Texas-Bexar County |
| California-Santa Clara County |
| New York-New York County |
| Michigan-Wayne County |
| California-Alameda County |
| Massachusetts-Middlesex County |
| Pennsylvania-Philadelphia County |
| Florida-Palm Beach County |
| California-Sacramento County |
| New York-Suffolk County |
| Florida-Hillsborough County |
| Florida-Orange County |
| New York-Nassau County |
| New York-Bronx County |
| Texas-Travis County |
| Ohio-Franklin County |
| Michigan-Oakland County |
| Pennsylvania-Allegheny County |
| Minnesota-Hennepin County |
| Ohio-Cuyahoga County |
| California-Contra Costa County |
| North Carolina-Wake County |
| Virginia-Fairfax County |
| Utah-Salt Lake County |
| North Carolina-Mecklenburg County |
| Georgia-Fulton County |
| Arizona-Pima County |
| Texas-Collin County |
| Maryland-Montgomery County |
| Florida-Pinellas County |
| Hawaii-Honolulu County |
| New York-Westchester County |
| Florida-Duval County |
| Missouri-St. Louis County |

**Supplementary table 2:**
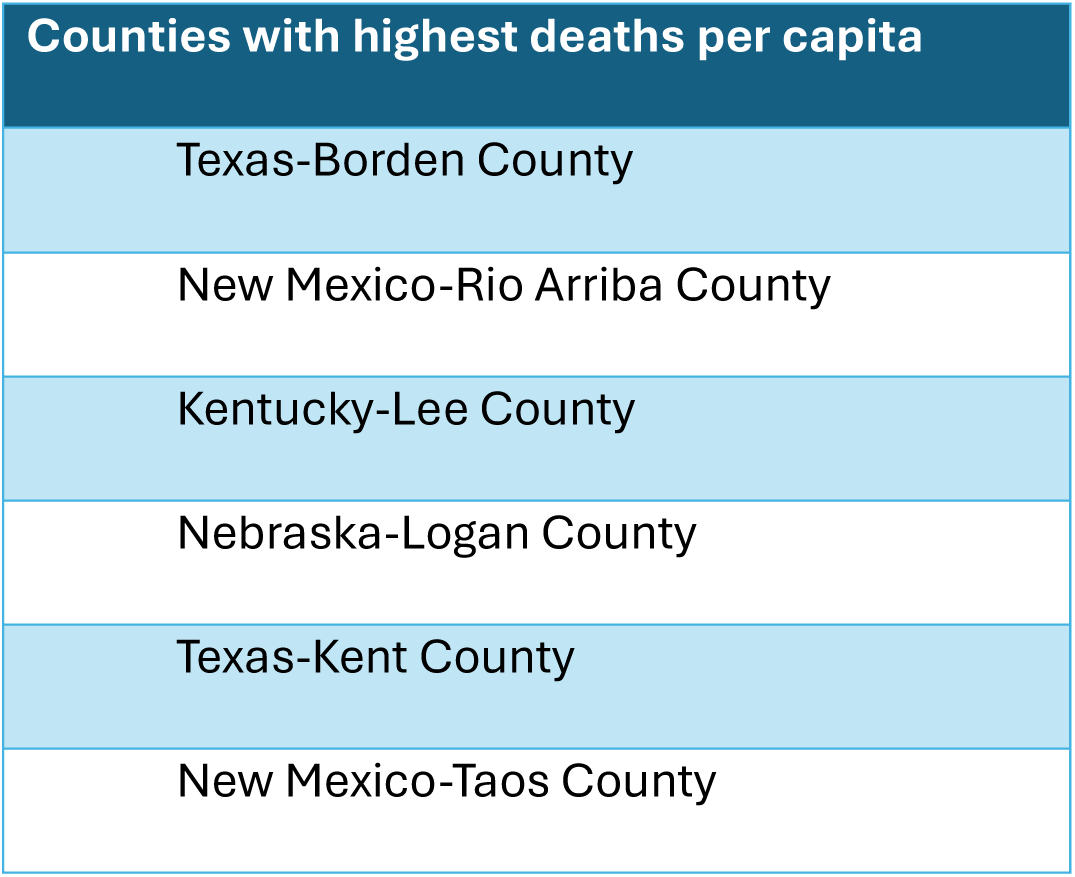
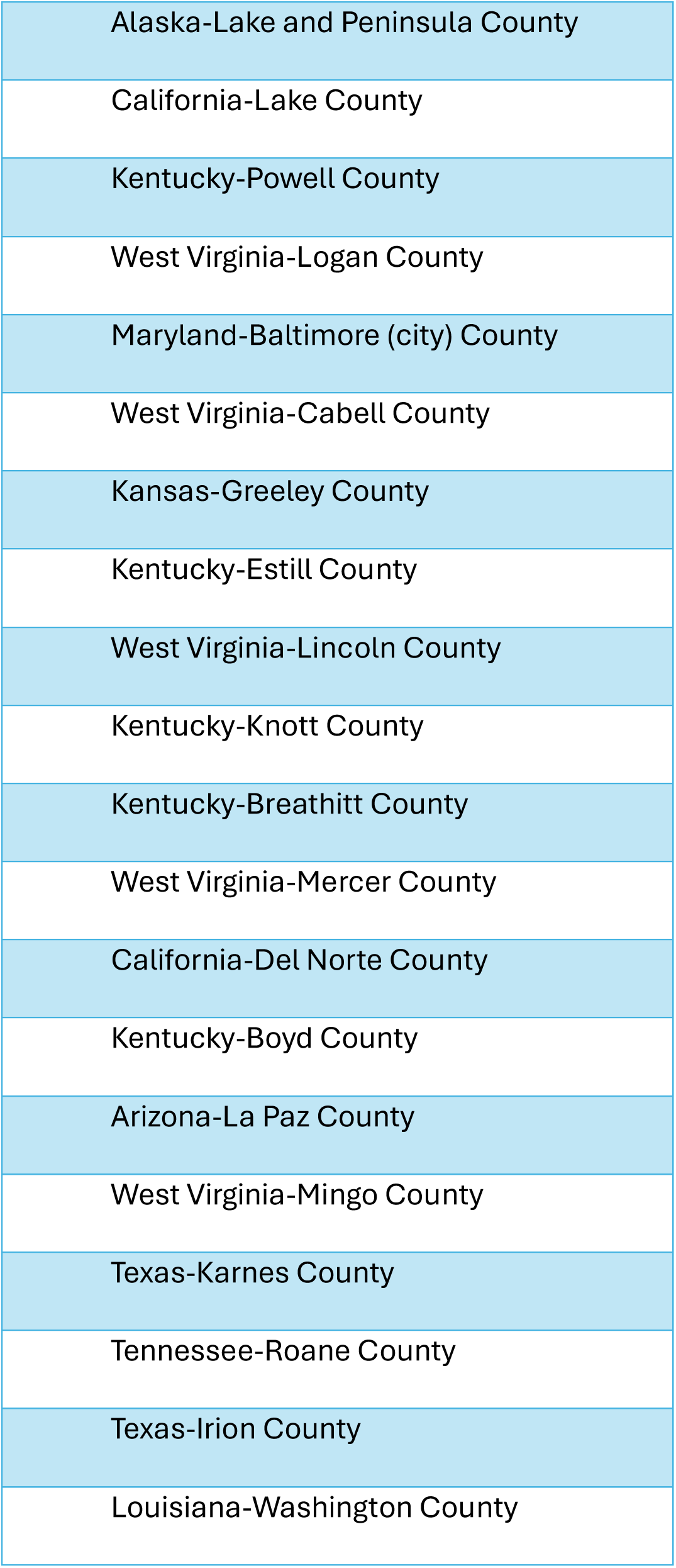
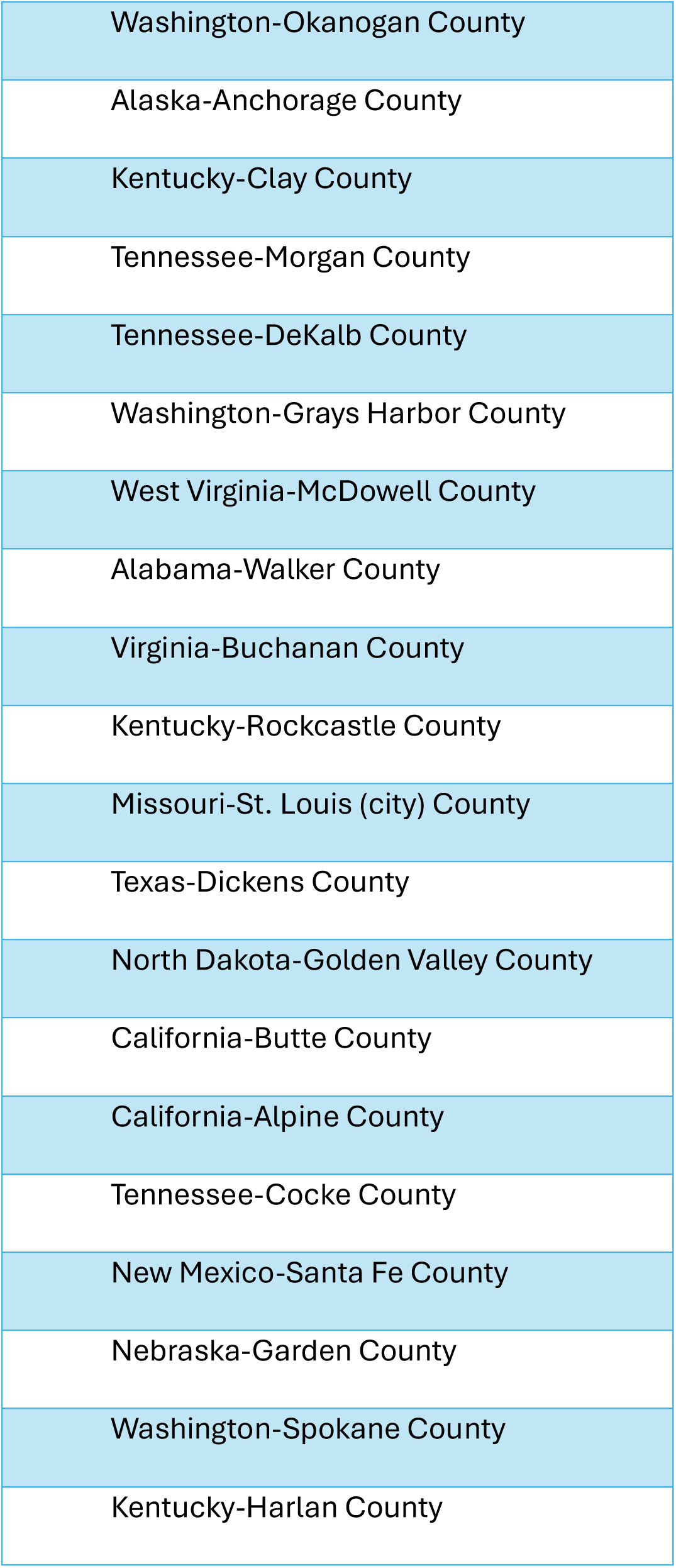
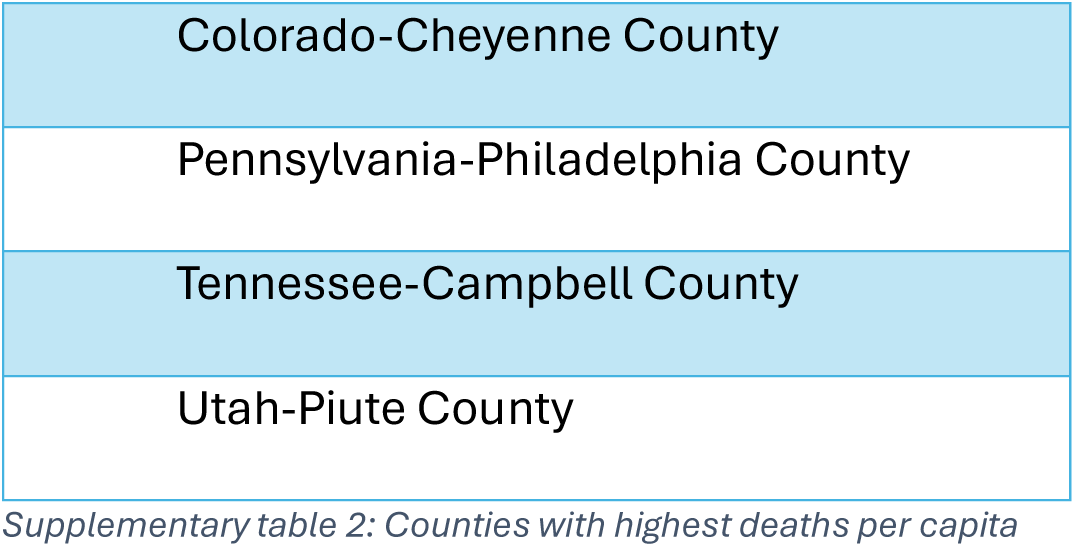
Counties with highest deaths per capita

